# A human brain network linked to restoration of consciousness after deep brain stimulation

**DOI:** 10.1101/2024.10.17.24314458

**Authors:** Aaron E.L Warren, Marina Raguž, Helen Friedrich, Frederic L.W.V.J. Schaper, Jordy Tasserie, Samuel B. Snider, Jian Li, Melissa M.J. Chua, Konstantin Butenko, Maximilian U. Friedrich, Rohan Jha, Juan E. Iglesias, Patrick W. Carney, David Fischer, Michael D. Fox, Aaron D. Boes, Brian L. Edlow, Andreas Horn, Darko Chudy, John D. Rolston

**Author notes:** Correspondence: Aaron E.L. Warren Hale Building for Transformative Medicine, 60 Fenwood Road, Boston, MA, USA. Joint senior authors (Darko Chudy, John D. Rolston).

## Abstract

Disorders of consciousness (DoC) are states of impaired arousal or awareness. Deep brain stimulation (DBS) is a potential treatment, but outcomes vary, possibly due to differences in patient characteristics, electrode placement, or stimulation of specific brain networks. We studied 40 patients with DoC who underwent DBS targeting the thalamic centromedian-parafascicular complex. Better-preserved gray matter, especially in the striatum, correlated with consciousness improvement.

Stimulation was most effective when electric fields extended into parafascicular and subparafascicular nuclei—ventral to the centromedian nucleus, near the midbrain— and when it engaged projection pathways of the ascending arousal network, including the hypothalamus, brainstem, and frontal lobe. Moreover, effective DBS sites were connected to networks similar to those underlying impaired consciousness due to generalized absence seizures and acquired lesions. These findings support the therapeutic potential of DBS for DoC, emphasizing the importance of precise targeting and revealing a broader link between effective DoC treatment and mechanisms underlying other conscciousness-impairing conditions.

## INTRODUCTION

There are no proven treatments for patients with chronic disorders of consciousness (DoC).^1, 2^ DoC are caused by brain injuries including hypoxia, ischemia, trauma, and intracerebral hemorrhage, resulting in impairments of arousal or awareness that vary widely in severity and prognosis.^3, 4^ These patients commonly reside in long-term care facilities with no or severely limited ability to engage with their environments—for months, years, even decades.

Neuromodulation has been explored as a potential therapy to restore consciousness in patients with DoC for over 50 years. Pioneering work by Hassler^5^ and McLardy^6^ in the 1960s was followed by larger case series of deep brain stimulation (DBS) in the 1990s^7, 8^ and beyond.^9–19^ Various stimulation targets have shown some efficacy in uncontrolled trials, including the intralaminar thalamus,^7–9, 12–14, 16, 17, 19^ brainstem,^8, 17^ pallidum,^5, 11^ and nucleus accumbens.^15^ However, evidence from randomized controlled trials is lacking.

Recent advances in understanding the brain networks underlying DoC^20^ have opened new avenues for diagnosis,^1, 2^ treatment,^21^ and outcome prediction.^4^ Regardless of the cause, DoC involve a widespread suppression of excitatory neurotransmission,^3^ particularly in the “mesocircuit”^20^ of the anterior forebrain, which includes the frontal cortex, central thalamus, striatum, and brainstem.

Within the thalamus, the posterior intralaminar nuclei (centromedian [CM] and parafascicular [Pf] nuclei) are central components of the mesocircuit^20^ and project directly to the striatum.^22, 23^ Based on these connections, the CM-Pf complex was targeted in a recent uncontrolled study of DBS for DoC,^9, 12^ the largest of its kind, motivated by findings in previous, smaller studies.^7, 8, 17, 18^ While no consistent effects were seen at the group level, this study identified a subset of patients with dramatic improvements in consciousness in the first year following implantation— beyond what has been seen in natural history studies of chronic DoC.^24, 25^ Here, we analyzed patient-level data to test whether these DBS “responders” integrity, that predicted treatment responsiveness. We also investigated whether they were stimulated in a specific thalamic subregion, white matter tract, or distributed functional brain network. Finally, we examined the external validity of this network by testing its involvement in separate groups of patients with lesion- or epileptic seizure-induced impairments of consciousness.

## RESULTS

### Patient characteristics and clinical outcomes

The analysis cohort included 40 patients with DoC secondary to cardiac arrest or traumatic brain injury who met criteria for DBS based on previously described clinical, neurophysiologic, and neuroimaging evaluations.^9, 12^ Patients were assessed using the Coma Recovery Scale-Revised (CRS-R)^26^ and classified as having unresponsive wakefulness syndrome (UWS), a minimally conscious state (MCS), or full consciousness.^1^ Supplementary clinical measures included the Disability Rating Scale (DRS) and Coma/Near-Coma (C/NC) scale.^27^

After a median of 6 months post-injury (IQR=3.5-13, range=2-137), patients underwent unilateral DBS targeting the left (*n=*37) or right (*n=*3) CM-Pf (**Fig. 1**). At 12 months post-DBS, 11/40 patients were classified as improved and 29 as non-improved. “Improved” patients were defined as those who transitioned from UWS to MCS or fully conscious, or from MCS to fully conscious, as in previous work.^9, 12^ Across all patients, the median increase in CRS-R scores was 2 (IQR=2-7, range=0-18; **Fig. 1**). Three patients died 3-6 months post-surgery. Secondary analyses were performed to test consistency of results when using a different outcome definition (each patient’s change in CRS-R scores, as opposed to a binary improved/non-improved classification), excluding patients with right-sided DBS implants, and excluding patients who died (see **Supplementary Material**).

**Figure 1:**
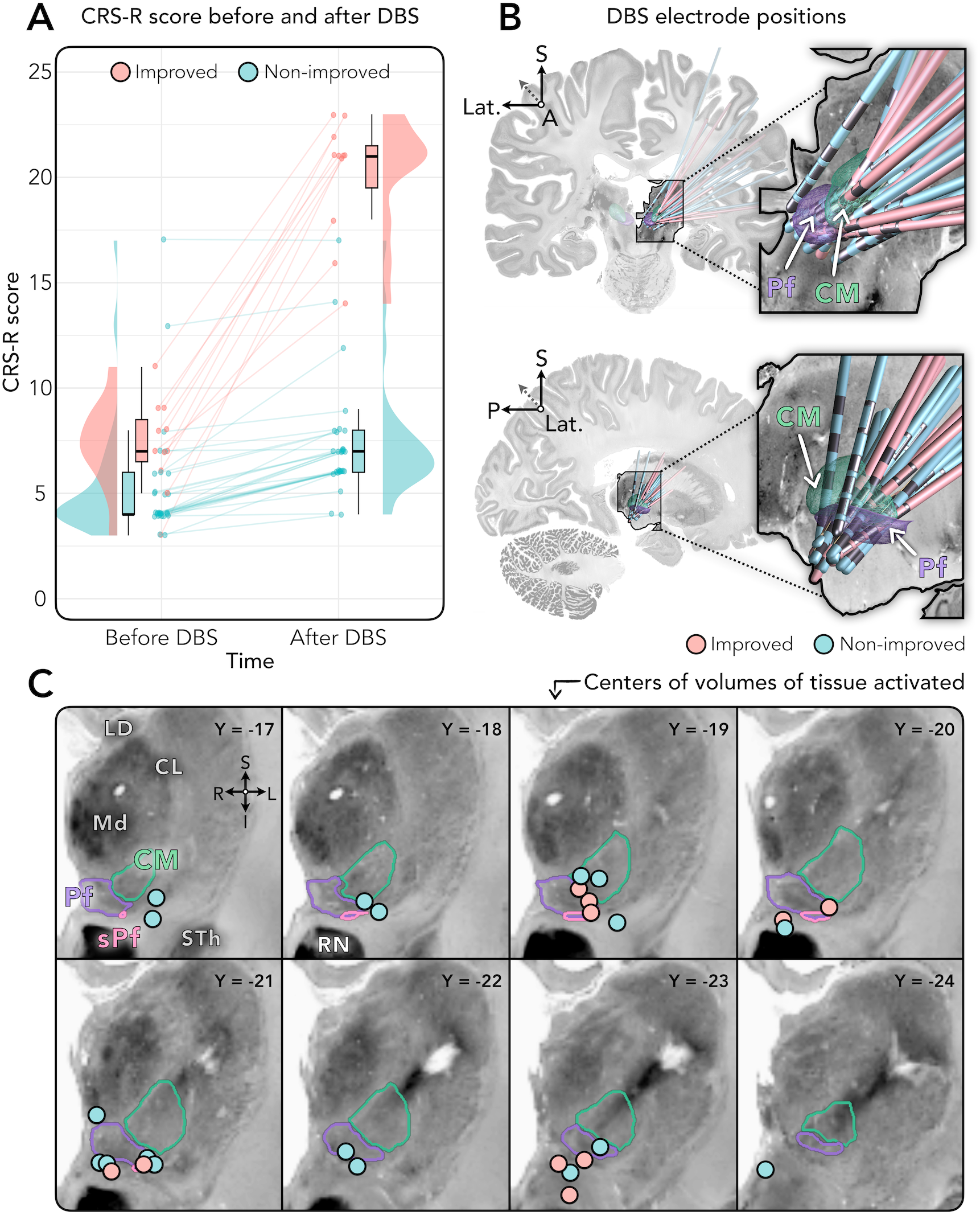
Clinical outcomes and DBS electrode localizations for the improved and non-improved patient groups. **(A)** Raincloud plots^28^ showing DoC severity measured before and 12 months after DBS using the Coma Recovery Scale-Revised (CRS-R), where higher scores correspond to better outcomes. Patients were categorized into improved (*n*=11) and non-improved (*n*=29) groups. **(B)** Three-dimensional DBS with respect to the centromedian (CM) and parafascicular (Pf) nuclei, as defined by the atlas of Krauth et al.^29^ based on the histology work of Morel.^30^ Leads are shown only for patients in whom accurate image registrations were possible (*n*=10 improved and *n*=18 non-improved). **(C)** Two-dimensional coronal views of the thalamus showing the center of each patient’s volume of tissue activated (i.e., the modelled electric field around DBS contact(s) chosen for stimulation). Pink and blue circles indicate locations for patients in the improved and non-improved groups, respectively. Y coordinates indicate the coronal position (in mm) of each slice, in MNI 152 ICBM 2009b nonlinear asymmetric template space. Stimulation coordinates for each patient are reported in **Supplementary Material**. Locations are displayed upon the BigBrain histological atlas^31^ registered to MNI space.^32^. <u>Abbreviations</u>: A, Anterior, CL, Central lateral nucleus, CM, Centromedian nucleus, CRS-R, Coma Recovery Scale—Revised, I, Inferior, L, Left, Lat., Lateral, LD, Lateral dorsal nucleus, Md, Mediodorsal nucleus, Pf, Parafascicular nucleus, R, Right, RN, Red nucleus, S, Superior, sPf, Subparafascicular nucleus, STh, Subthalamic nucleus.

We first tested the hypothesis that clinical variables are associated with improvement (**Table 1**). Compared to non-improved patients, those who improved had less severe baseline impairments on the DRS and C/NC scales (*p*<0.05, false of 20 years younger at the time of injury. In contrast, the groups did not differ by sex, side of DBS, etiology of DoC, duration between initial injury and DBS, DBS lead model, or the programmed stimulation amplitude, frequency, or pulse width.

**Table 1:**
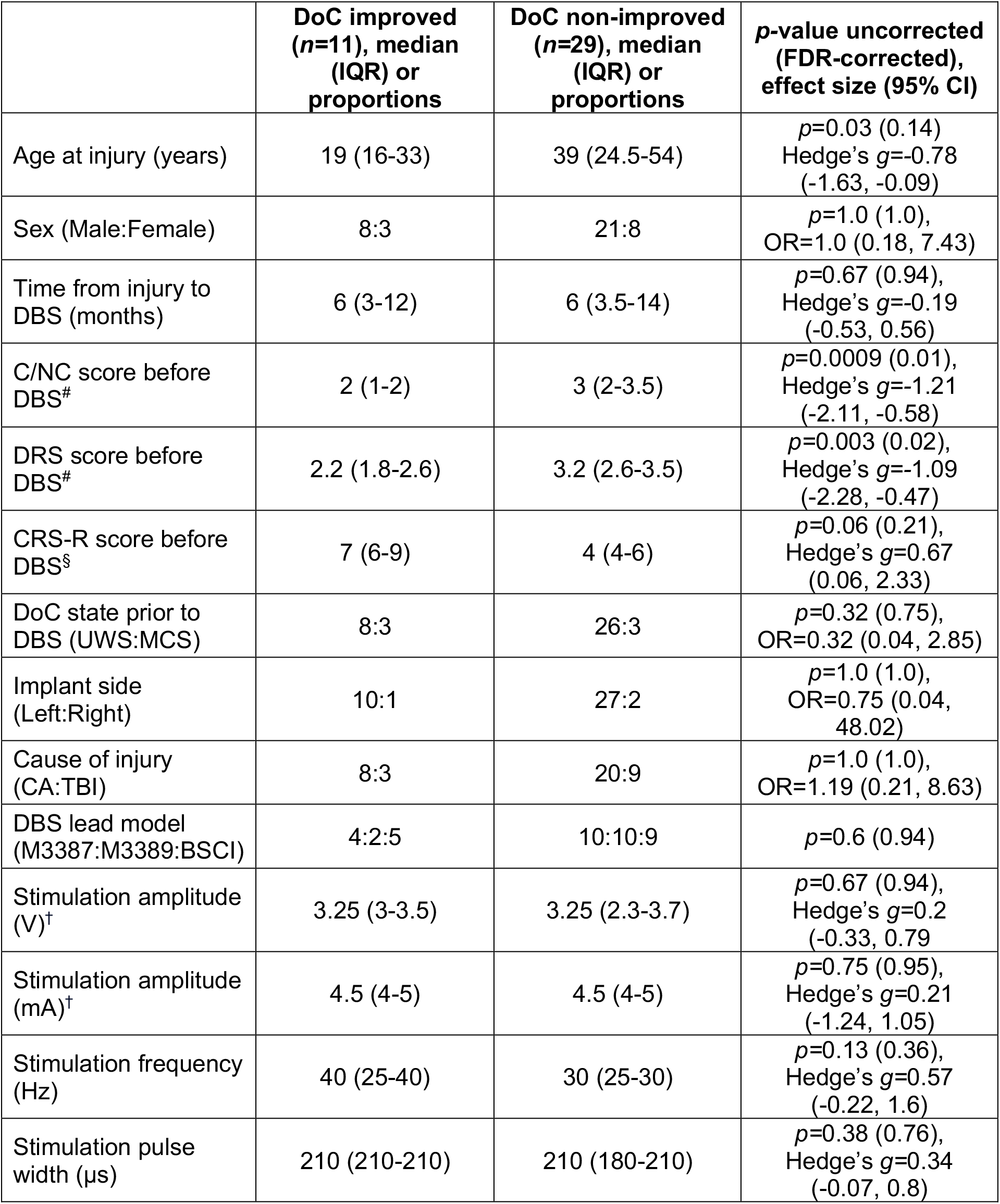
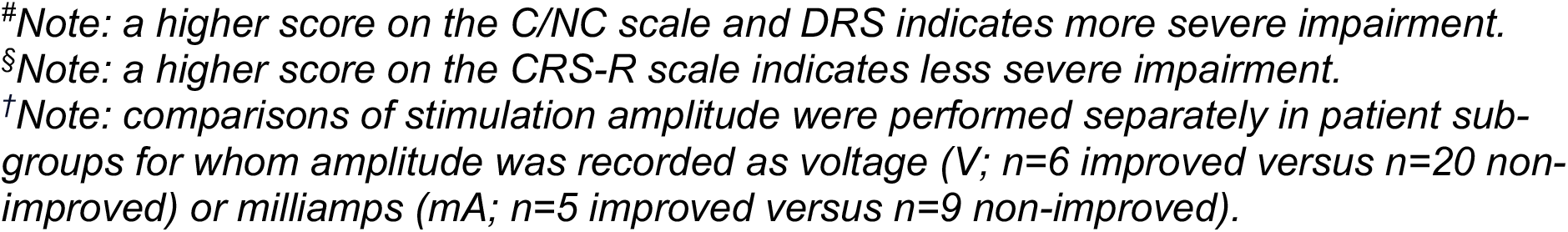
Comparison of demographic and clinical variables between patients who improved (*n*=11) and those who did not improve (*n*=29) with DBS. Results are reported using uncorrected and false discovery rate (FDR)-corrected p-values, the latter corrected for 14 clinical and demographic variables tested. *<u>Abbreviations</u>*: BSCI, Boston Scientific Vercise lead model; CA, Cardiac arrest; CI, Confidence interval; CNC, Coma/near-coma; M3387, Medtronic lead model 3387; DRS, Disability Rating Scale; TBI, Traumatic brain injury; UWS, Unresponsive wakefulness syndrome.

|  | DoC improved<br>(*n*=11), median<br>(IQR) or<br>proportions | DoC non-improved<br>(*n*=29), median<br>(IQR) or<br>proportions | *p*-value uncorrected<br>(FDR-corrected),<br>effect size (95% CI) |
| --- | --- | --- | --- |
| Age at injury (years) | 19 (16-33) | 39 (24.5-54) | *p*=0.03 (0.14)<br>Hedge's *g*=-0.78<br>(-1.63, -0.09) |
| Sex (Male:Female) | 8:3 | 21:8 | *p*=1.0 (1.0),<br>OR=1.0 (0.18, 7.43) |
| Time from injury to<br>DBS (months) | 6 (3-12) | 6 (3.5-14) | *p*=0.67 (0.94),<br>Hedge's *g*=-0.19<br>(-0.53, 0.56) |
| C/NC score before<br>DBS<sup>#</sup> | 2 (1-2) | 3 (2-3.5) | *p*=0.0009 (0.01),<br>Hedge's *g*=-1.21<br>(-2.11, -0.58) |
| DRS score before<br>DBS<sup>#</sup> | 2.2 (1.8-2.6) | 3.2 (2.6-3.5) | *p*=0.003 (0.02),<br>Hedge's *g*=-1.09<br>(-2.28, -0.47) |
| CRS-R score before<br>DBS<sup>§</sup> | 7 (6-9) | 4 (4-6) | *p*=0.06 (0.21),<br>Hedge's *g*=0.67<br>(0.06, 2.33) |
| DoC state prior to<br>DBS (UWS:MCS) | 8:3 | 26:3 | *p*=0.32 (0.75),<br>OR=0.32 (0.04, 2.85) |
| Implant side<br>(Left:Right) | 10:1 | 27:2 | *p*=1.0 (1.0),<br>OR=0.75 (0.04,<br>48.02) |
| Cause of injury<br>(CA:TBI) | 8:3 | 20:9 | *p*=1.0 (1.0),<br>OR=1.19 (0.21, 8.63) |
| DBS lead model<br>(M3387:M3389:BSCI) | 4:2:5 | 10:10:9 | *p*=0.6 (0.94) |
| Stimulation amplitude<br>(V)<sup>†</sup> | 3.25 (3-3.5) | 3.25 (2.3-3.7) | *p*=0.67 (0.94),<br>Hedge's *g*=0.2<br>(-0.33, 0.79) |
| Stimulation amplitude<br>(mA)<sup>†</sup> | 4.5 (4-5) | 4.5 (4-5) | *p*=0.75 (0.95),<br>Hedge's *g*=0.21<br>(-1.24, 1.05) |
| Stimulation frequency<br>(Hz) | 40 (25-40) | 30 (25-30) | *p*=0.13 (0.36),<br>Hedge's *g*=0.57<br>(-0.22, 1.6) |
| Stimulation pulse<br>width (μs) | 210 (210-210) | 210 (180-210) | *p*=0.38 (0.76),<br>Hedge's *g*=0.34<br>(-0.07, 0.8) |

*#Note: a higher score on the C/NC scale and DRS indicates more severe impairment.*
*§Note: a higher score on the CRS-R scale indicates less severe impairment.*
*†Note: comparisons of stimulation amplitude were performed separately in patient sub-* *groups for whom amplitude was recorded as voltage (V; n=6 improved versus n=20 non-* *improved) or milliamps (mA; n=5 improved versus n=9 non-improved).*

### Optimal brain tissue integrity

We next investigated whether DBS outcomes were associated with MRI measures of brain tissue integrity (**Fig. 2**) in patients with T1-weighted MRI available (*n=*8 improved patients versus *n=*18 non-improved). MRI scans were segmented into whole-brain (gray matter, white matter, cerebrospinal fluid) and regional subcortical volumes,^33^ then normalized by total intracranial volume and by age-matched controls.^34^ At the uncorrected significance threshold, patients who improved had greater preservation of whole-brain gray matter and larger volumes of cerebellar gray matter and the dominant striatal projections of the CM-Pf, including the putamen and caudate. These comparisons did not survive FDR correction. The groups did not differ in thalamic, pallidal, or brainstem volumes.

**Figure 2:**
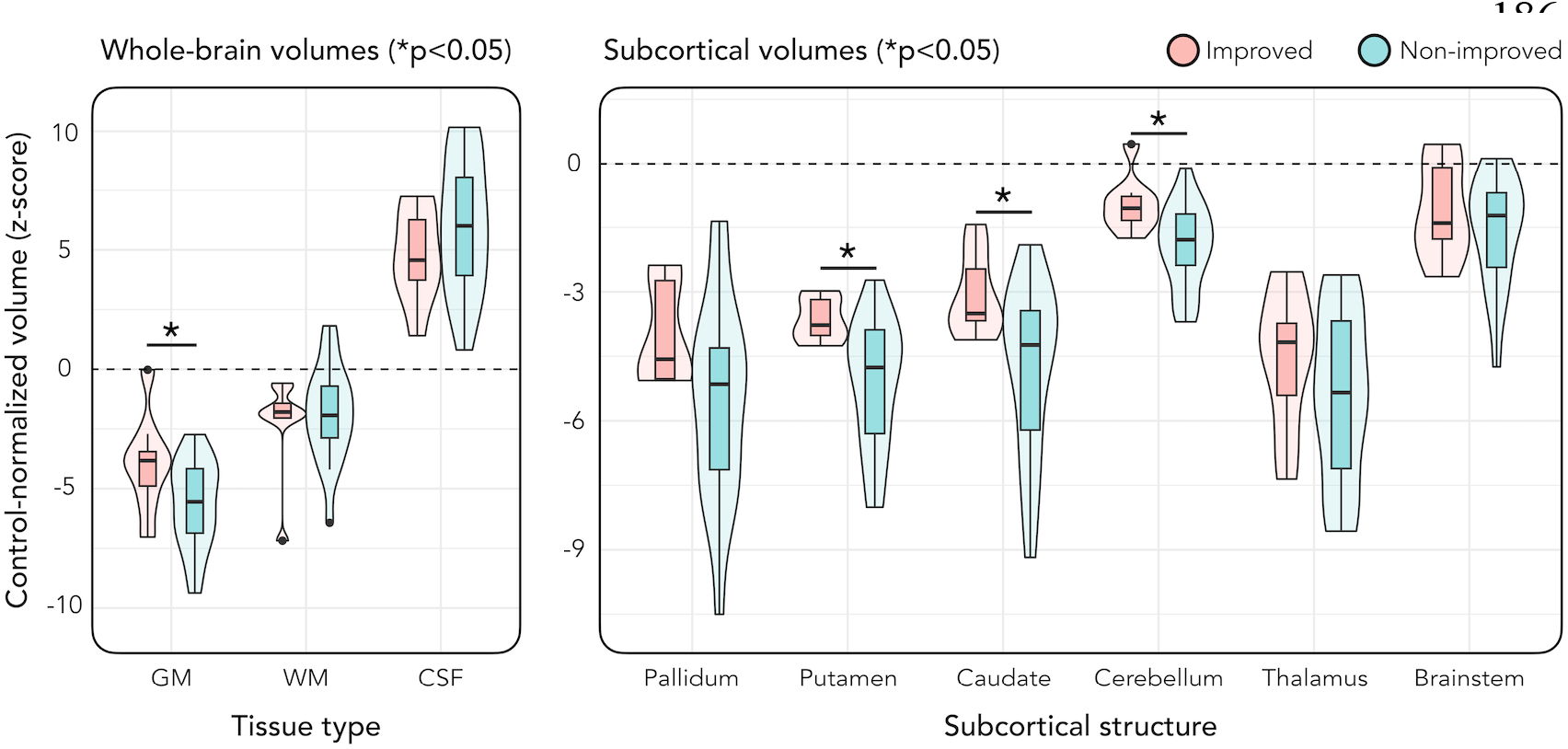
Comparison of MRI tissue volumes between improved and non-improved groups. Violin plots showing whole-brain volumes of gray matter (GM), white matter (WM), and cerebrospinal fluid (CSF) in the improved (*n=*8) and non-improved (*n=*18) groups. This analysis only included patients with T1-weighted MRI available. The plot on the right shows comparisons of subcortical gray matter volumes. The dashed horizonal line on each plot indicates the average value in age-matched controls from the Nathan Kline Institute-Rockland Sample (NKI-RS).^34^ Volumes from patients are expressed as z-scores measuring the distance, in units of SD, away from the control mean. * = *p<*0.05 (uncorrected).

### Optimal stimulation site

Electrodes were localized using Lead-DBS software.^35^ This revealed variability in electrode placement across patients (**Fig. 1**), suggesting outcomes may be linked to stimulation of specific thalamic subregions. To test this, we calculated electric fields (E-fields) for each patient, which estimate the distribution and magnitude of stimulation based on the DBS settings.^35, 36^ Then, to identify sites linked to therapeutic benefit (**Fig. 3**), we initially conducted a series of voxel-wise two-sample *t-*tests to pinpoint potential sites of optimal stimulation (“sweet spots”).

**Figure 3:**
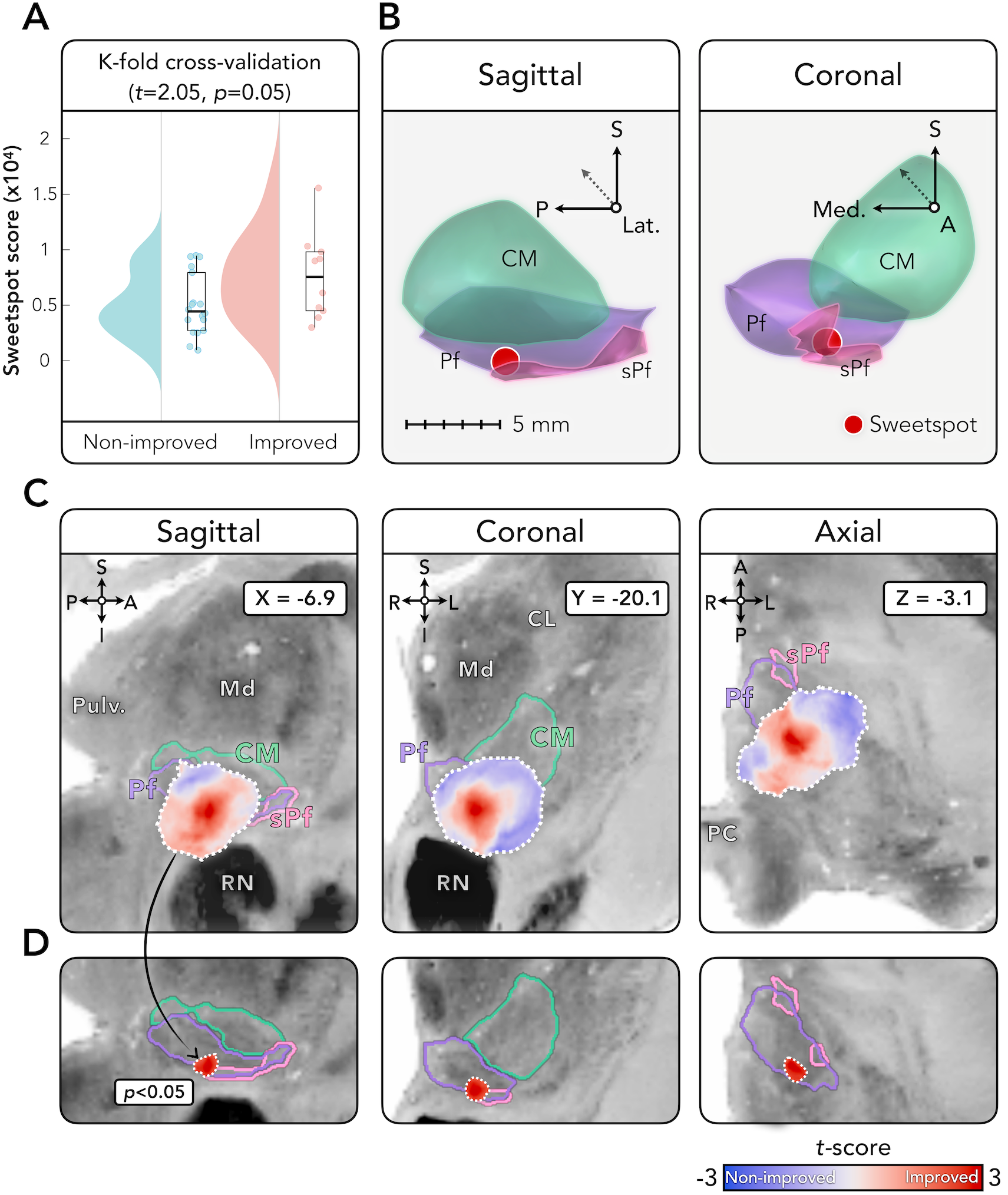
Anatomical localization and cross-validation of optimal stimulation site. **(A)** K-fold (*k*=10) cross-validation showing that E-field peak locations are associated with clinical outcomes in left-out patients (*p*=0.05). **(B)** Three-dimensional views displaying the location of the stimulation “sweet spot”, defined as the center-of-gravity of the largest cluster (*p*<0.05, uncorrected) following voxel-wise two-sample t-tests of E-field magnitudes between improved (*n*=10) and non-improved (*n*=18) groups. The sweet spot is shown in sagittal and coronal orientations with respect to the centromedian (CM), parafascicular (Pf), and subparafascicular (sPf) thalamic nuclei, as defined by the atlas of Krauth et al.^29^ based on the histology work of Morel.^30^ **(C)** Two-dimensional views of the thalamus showing an unthresholded map of *t*-scores where positive values indicate locations where E-field magnitudes were higher in the improved relative to non-improved group, and negative values where Thresholded map (*p*<0.05, uncorrected) showing the peak site associated with therapeutic benefit. Results are displayed upon the BigBrain histological atlas^31^ registered to MNI space.^32^ <u>Abbreviations</u>: A, Anterior, CL, Central lateral nucleus, I, Inferior, L, Left, Lat., Lateral, Md, Mediodorsal nucleus, Med., Medial, P, Posterior, PC, Posterior commissure, Pulv, Pulvinar nucleus, R, Right, RN, Red nucleus, S, Superior.

This analysis compared E-field magnitudes between improved (*n=*10) and non-improved (*n=*18) groups. The *t-*tests identified a candidate sweet spot defined by the largest surviving voxel cluster at a significance threshold of *p*<0.05. This site was in the inferior Pf, aligned with the axial anterior commissure-posterior commissure (AC-PC) line, and extended into the subparafascicular nucleus below, with MNI 152 ICBM 2009b coordinates (mm) of [*X*=-6.91, *Y*=-20.11, *Z*=- 3.08]. Notably, there were no regions where the E-field magnitude was higher in the non-improved group. To test the reliability of this candidate site, we subjected the results to *k-*fold cross-validation. This confirmed the robustness of the findings (*t*=2.05, *p*=0.05), validating the candidate sweet spot’s association with benefit.

### Optimal structural connectivity

Having identified optimal DBS sites, we next hypothesized that DBS improvement might be mediated by structural connections traversing areas of beneficial stimulation.^35^ To test this, we calculated white matter streamlines using a normative diffusion MRI connectome acquired at an ultra-high resolution of 760 µm,^37^ as in our recent work.^38, 39^ Like the “sweet spot” analysis, which analyzed stimulations across voxels, here we compared stimulation magnitudes across *streamlines* between improved (*n=*10) and non-improved (*n=*18) groups. This involved conducting similar two-sample *t-*tests to pinpoint potential streamlines associated with therapeutic benefit, at an initial threshold of *p*<0.05. Improved patients had stronger involvement of connections to distributed projection pathways of the ascending arousal network,^40^ including the hypothalamus (posterior, periventricular, and paraventricular hypothalamic nuclei), midbrain (red nucleus, mesencephalic reticular formation, and ventral tegmental area), pons (locus coeruleus, subcoeruleus, and medial parabrachial nucleus), medulla (inferior and superior medullary reticular formation), cerebellar dentate nucleus, and medial frontal cortex (**Fig. 4**). There were no connections with stronger involvement in non-improved patients. The findings were robust to *k-*fold cross-validation testing (*t=*2.29, *p*=0.03), confirming their association with benefit.

**Figure 4:**
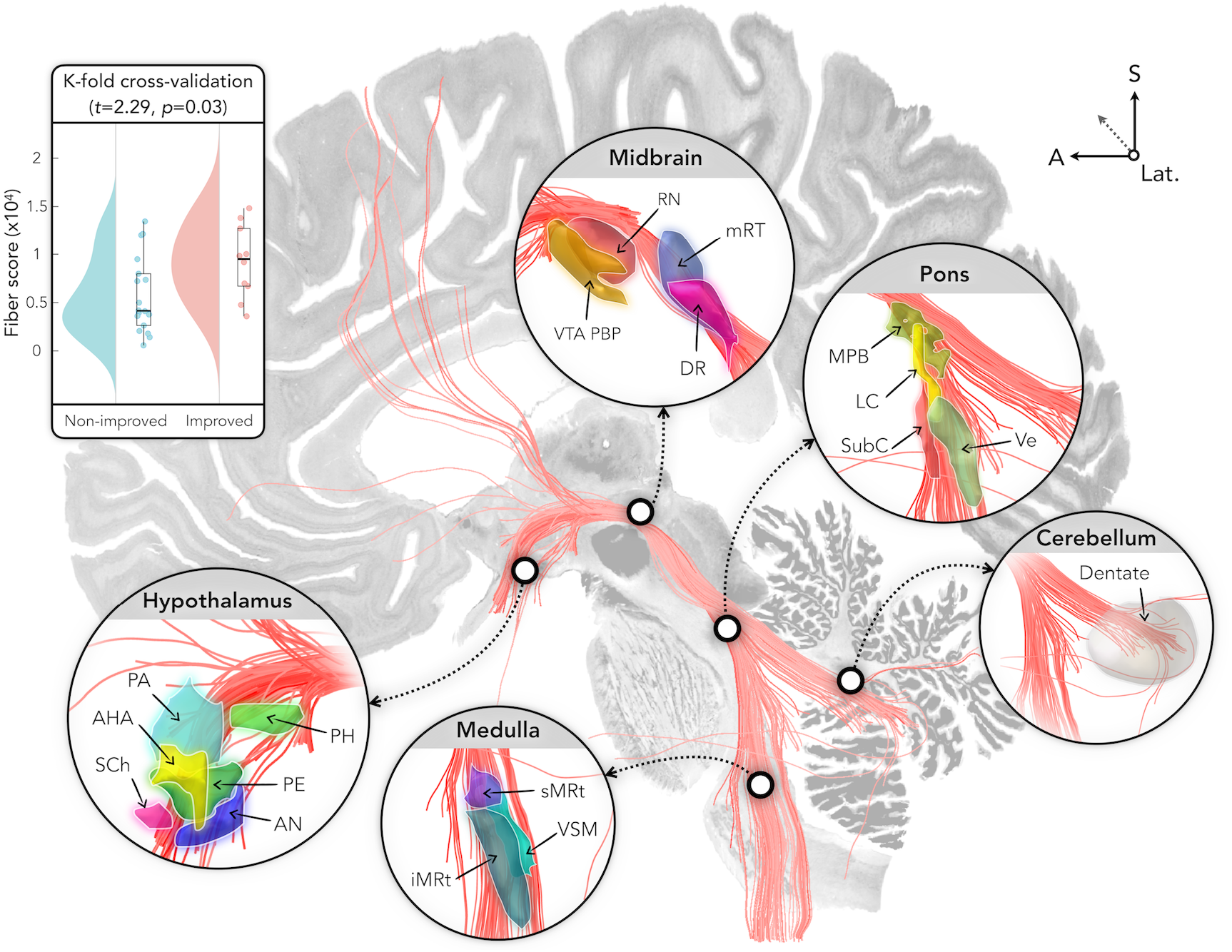
Anatomical localization and cross-validation of optimal structural connectivity. White matter fiber tracts more strongly involved in the improved (*n*=10) relative to non-improved (*n*=18) group (*p*<0.05, uncorrected). Box plots in the upper left corner show the results of K-fold (*k*=10) cross-validation, demonstrating that fiber tracts are associated with clinical outcome in left-out patients (*p*=0.03). The most strongly connected subcortical nuclei are displayed using published atlases of the hypothalamus^41^ (https://zenodo.org/records/3942115), brainstem (https://www.nitrc.org/projects/brainstemnavig), ascending arousal network^40^ (https://doi.org/10.5061/dryad.zw3r228d2), and cerebellum^42^ (https://www.diedrichsenlab.org/imaging/propatlas.htm). Results are displayed upon the BigBrain histological atlas^31^ registered to MNI space.^32^ <u>Abbreviations</u>: A, Anterior, AHA, Anterior hypothalamic area, AN, Arcuate nucleus, DR, Dorsal raphe nucleus, iMRt, Inferior medullary reticular formation, Lat., Lateral, LC, Locus coeruleus, MPB, Medial parabrachial nucleus, mRt, Mesencephalic reticular formation, PA, Paraventricular nucleus, PE, Periventricular nucleus, PH, Posterior hypothalamus, RN, Red nucleus, S, Superior, SCh, Suprachiasmatic nucleus, sMRt, Superior medullary reticular formation, SubC, Subcoeruleus, Ve, Vestibular nuclei complex, VSM, Viscero-sensory-motor nuclei complex, VTA PBP, Ventral tegmental area (parabrachial pigmented nucleus complex).

### Optimal functional connectivity

In the next analysis, we investigated blood-oxygen-level-dependent (BOLD) connectivity of DBS sites using a normative resting-state fMRI dataset acquired in normative fMRI data and calculating connectivity with all brain voxels. Connectivity strengths were then compared between improved (*n=*10) and non-improved (*n=*18) groups using voxel-wise two-sample *t-*tests, resulting in a map where positive *t-*scores indicate higher connectivity in the improved group (**Fig. 5A**).

**Figure 5:**
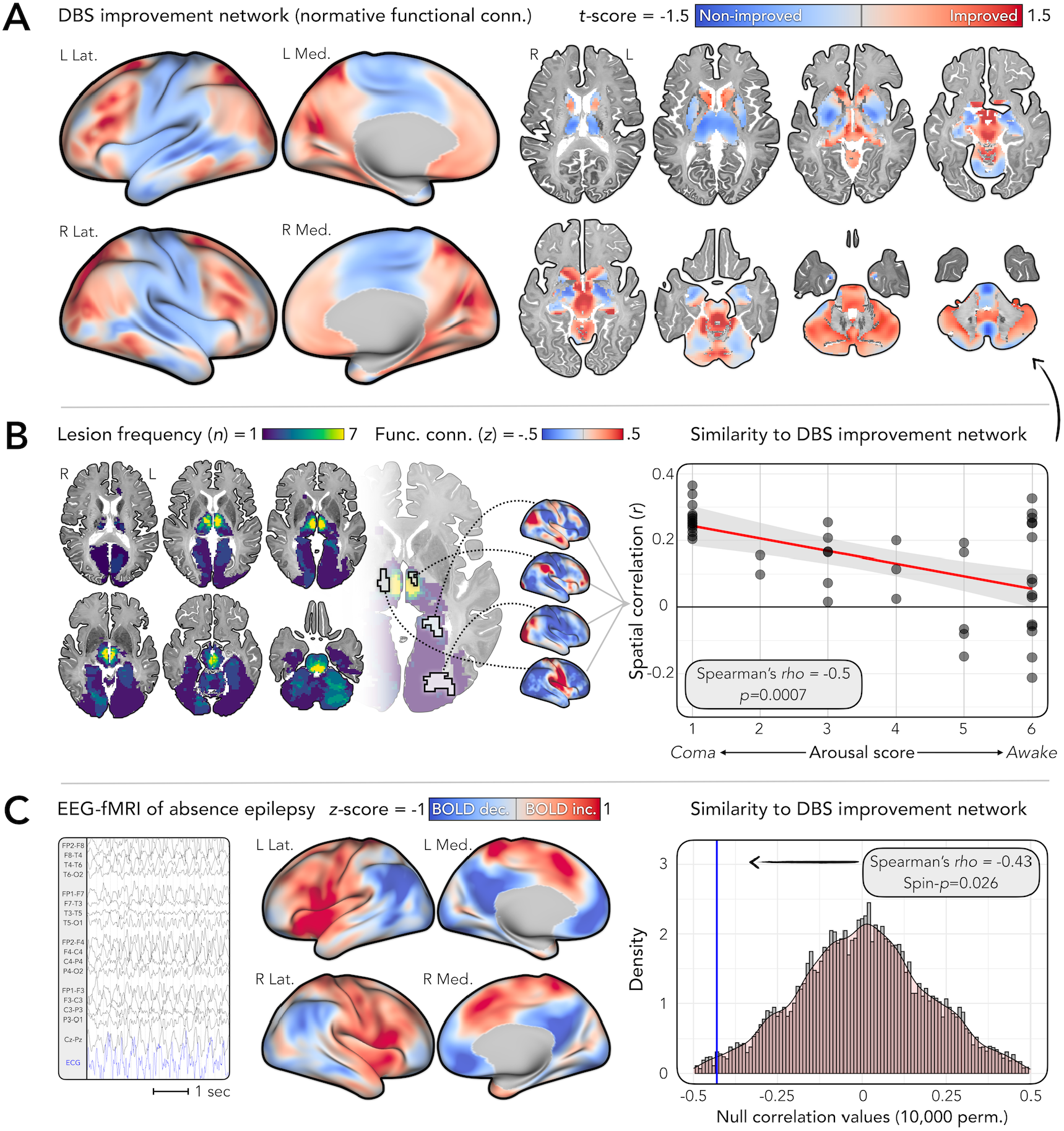
Optimal functional connectivity and alignment with brain networks disrupted in other consciousness-impairing conditions. **(A)** A DBS improvement network was calculated by comparing functional connectivity of stimulation sites between improved *(n*=10) and non-improved (*n*=18) groups, using normative fMRI data.^43, 44^ Results are *t*-scores (unthresholded), where positive values indicate regions of higher connectivity in the improved group and negative values indicate lower connectivity. Cortical views are shown using the *fs_LR_32k* template (https://balsa.wustl.edu/QXj2) and subcortical views using the BigBrain histological atlas^31^ registered to MNI space.^32^. **(B)** We spatially compared the DBS improvement network (from A) to patterns of functional connectivity seen in a separate group of 45 patients with acute-onset, arousal-impairing lesions.^45, 46^ Axial views are frequency maps showing all lesion locations. For each lesion, we calculated a whole-brain functional connectivity map. We then assessed spatial similarity between each map and our DBS improvement network. The scatter plot shows a significantly negative association with arousal outcomes in the lesional group (measured as a 6-point rating, where lower values indicate more severe impairment; for details, see^45, 46^). **(C)** We also assessed level-dependent (BOLD) signal change during generalized spike-wave discharges in 15 patients with absence epilepsy scanned using simultaneous EEG with functional MRI (EEG-fMRI).^48–50^ The EEG trace shows an example of these discharges recorded inside the MRI scanner (for methodological details, refer to^48–50^). EEG-fMRI results are displayed as group-level *z*-scores where positive values indicate increased BOLD signal during discharges and negative values indicate decreased BOLD signal. The histogram/density plot shows the results of spin permutation testing (10,000 spins),^51^ demonstrating a significantly negative correlation between the EEG-fMRI map and the DBS improvement network. <u>Abbreviations</u>: Conn., Connectivity, EEG, Electroencephalogram, ECG, Electrocardiogram, Func., Functional, L, Left, R, Right.

The primary purpose of this analysis was to calculate a whole-brain, spatially continuous (i.e., unthresholded) map for subsequent comparison with external datasets of patients with consciousness-impairing brain lesions and seizures (see below). To identify the cortical networks most implicated in improvement, we calculated the mean *t-*score within each of the seven canonical “Yeo atlas” networks.^43^ Networks showing a positive mean *t-*score (i.e., more associated with improvement) were, in descending order, the visual, dorsal attention, frontoparietal, and default-mode networks. In contrast, the somatomotor and ventral attention networks showed a negative mean *t-*score (non-improvement).

Applying a voxel-wise threshold of *p*<0.05 (uncorrected), the improved group showed stronger connectivity with the hypothalamus, midbrain, pons, dorsal cerebellum, posterior hippocampus, and parieto-occipital fissure (**Fig. 5A**).

### Alignment with consciousness-impairing lesion network

Our previous analyses aimed to identify the brain network underlying favorable DBS outcomes. To assess the external validity of these findings, we tested whether effective DBS for DoC modulates the same networks disrupted by brain lesions that cause impaired arousal in a separate cohort. Specifically, we tested whether the DBS improvement network (**Fig. 5A**) overlapped the pathological brain networks in 45 patients who had acute thalamic or brainstem lesions from stroke or trauma. As previously described,^45, 46^ these patients were ordinally ranked based on the severity of their arousal impairment post-injury using scores from 1-6, with lower scores indicating more severe impairment. For each patient, we examined the brain network connected to their lesion using the same normative fMRI dataset described earlier,^43, 44^ producing 45 lesion connectivity maps, each linked to a patient’s arousal score. We then calculated the spatial similarity (Pearson correlation) between these lesion connectivity maps and our worse outcomes (lower arousal scores). Consistent with this, we found that lesions associated with worse arousal impairments were connected to the positive regions in our DBS improvement network (Spearman *rho*=-0.5, *p=*0.0007; **Fig. 5B**). Similarly, when restricting the analysis to patients with coma (*n=*14) and those who were awake (*n=*15),^45, 46^ we found that coma patients’ lesion connectivity maps had higher similarity to our DBS improvement network than awake patients (*t=*3.8, *p=*0.0006, permutation-based two-sample *t-*test).

### Alignment with consciousness-impairing seizure network

In a final analysis, we tested whether effective DBS for DoC modulates the same network disrupted by absence seizures, which are brief lapses of awareness marked by generalized spike-wave discharges on scalp EEG.^47^ We used findings from a previous study of 15 patients with absence epilepsy who underwent simultaneous EEG-fMRI,^48–50^ a technique that can measure whole-brain BOLD signal changes time-locked to epileptiform EEG events. This analysis produced a group-level brain map showing areas linked to absence seizure-related disruption of awareness (**Fig. 5C**). Like in our earlier analysis of arousal-impairing brain lesions, we aimed to compare this EEG-fMRI map with our DBS improvement network, hypothesizing that the brain network where DBS improves consciousnesses overlaps with the network where seizures disrupt it. Using spin-permutation testing,^51^ we found that areas of BOLD signal suppression during generalized spike-wave discharges overlapped the positive regions in our DBS improvement network (Spearman *rho=*-0.43, *p=*0.026); this BOLD suppression is thought to contribute to the transient lapses of awareness during absence seizures^47, 52^ and is most prominent in areas of the default-mode network.^48–50^

### Secondary analyses

Results were similar across three secondary analysis designs: (i) using a finer outcome measure based on the change in total CRS-R scores (instead of a binary improved/non-improved classification), (ii) excluding three patients who died within 12 months of DBS, and (iii) excluding three patients with right- (instead of left-) sided DBS implants (**Supplementary Material**). Specifically, all secondary analyses showed that improvement was linked to less severe baseline impairment (DRS and C/NC scores), younger age at injury, and larger volumes of the putamen and cerebellum (*p*<0.05, uncorrected). Patients who improved also continued to show larger volumes of whole-brain gray matter and the caudate when using CRS-R scores and when excluding patients who died (*p*<0.05, uncorrected), but not when excluding patients with right-sided implants (*p*>0.05).

Regarding optimal structural connectivity, the fiber tract distributions linked to DBS improvement were like those in the primary analysis; the findings remained robust to *k-*fold cross-validation when using CRS-R scores (Pearson *r*=0.42, *p*=0.028), and when excluding patients with right-sided implants (*t=*2.29, *p*=0.03), but did not reach significance when excluding patients who died (*t=*1.18, *p*=0.25). Finally, for optimal stimulation sites, the peak location associated with improvement remained in the ventral parafascicular nucleus, with a Euclidean distance of <0.7 mm from the primary analysis peak in all secondary designs. However, unlike the primary analysis, these secondary analyses did not survive cross-validation (*p>*0.05).

## DISCUSSION

We investigated the optimal preoperative clinical features, brain tissue integrity, stimulation target, and connectivity for CM-Pf DBS in DoC. Our findings suggest the importance of selecting patients based on their latent capacity for recovery and neuroplasticity^13, 53^ and stimulation targets based on their position and connectivity within the ascending arousal network.^40, 54–56^ The results shed light on potential mechanisms through which disparate stimulation targets trialed over the past 50 years may exert therapeutic effects for DoC,^5–19^ and provide foundational data to design future prospective and controlled trials.

Patients who improved with DBS were younger and had less severe baseline impairment. These findings align with prior studies on the influence of age and cognitive reserve on recovery after brain injuries.^13, 19, 57^ In stroke^58^ and TBI,^59^ older patients show slower response to rehabilitation than younger patients with comparable injuries. Similar trends occur in DBS for Parkinson’s disease^19^ and transcranial magnetic stimulation for depression,^60^ where younger age predicts better outcomes (though conflicting findings exist).^61^ This may reflect reduced capacity for brain plasticity and repair^62^ in older age, limiting stimulation-induced changes in neurogenesis,^63^ myelination,^64^ gene expression,^65, 66^ and neurotransmitter release,^67^ all of which may contribute to DoC recovery.^3, 21^

Improvement was seen in patients with more intact gray matter volumes on structural MRI, at the whole-brain level and more specifically in the striatum and cerebellum. Like younger age, gray matter preservation may be essential for treatment effects of DBS to take hold, whether in the short-term (i.e., acute functional changes requiring intact neuronal assemblies, like the synaptic transmission of stimulation) or longer term (i.e., chronic functional and/or structural neuroplastic changes).^68, 69^ In keeping with this hypothesis, we previously found that effective CM-Pf DBS for DoC is associated with longitudinal gray matter volume increases up to seven years post-DBS, including in the striatum.^70^

Preservation of the striatum and improved capacity for recovery is consistent with from a diffuse suppression of synaptic input from the cortex to striatal medium spiny neurons, leading to a loss of inhibitory projections to the global pallidus and in turn a tonic inhibition of the thalamus, culminating in a breakdown of anterior forebrain arousal.^20^ Increasing activity within this circuit, for example by DBS, is thus thought to restore consciousness. Preserved volume in improved patients suggests the striatum may play a gating role^71^ in restoring mesocircuit activity and thus could serve as a preoperative marker to identify optimal DBS candidates.

Despite targeting the CM-Pf region in all patients, there was natural variability in electrode positions and stimulation. Correlating this variability with outcomes, we found that improved patients had greater stimulation of the inferior Pf and subparafascicular nucleus. Although the CM and Pf are often described as a unitary complex, the two nuclei can be distinguished anatomically,^72^ functionally,^73, 74^ and connectomically,^22, 23, 75^ which may confer differential stimulation effects. In primates, the CM and Pf are dominant sources of glutamatergic input to the striatum, but their projection profiles differ: the CM projects to striatal territories receiving input from sensorimotor cortex, particularly the caudal putamen, while the Pf projects to association and limbic territories, including the anterior putamen, caudate, and nucleus accumbens.^22, 23, 75^ Their extra-striatal projections also differ, with the Pf having more input to the hypothalamus, amygdala, and ventral tegmental area, and to prefrontal, anterior cingulate, and frontal eye field regions.^22, 23^ Among the posterior intralaminar nuclei, the subparafascicular nucleus has the densest descending projections to the brainstem, including the inferior olivary nucleus, peripeduncular area, reticular core, and raphe nuclei.^23^

The CM is a more recent evolutionary development, with maximal expansion in primates. In smaller-brained species (e.g., rodents), the CM is not clearly distinct from the Pf.^72^ This suggests that the Pf may have a more conserved role in arousal, a function of all vertebrate brains, even those lacking a distinct CM.

Hence, one explanation for the sweet spot being in the Pf and subparafascicular nucleus stems from the preferential effects that DBS of this region may have upon associative and limbic areas of the striatum and cortex, and the resulting influence including striatal inputs to the global pallidus (potentially counteracting abnormal inhibition of the thalamus)^20^ and frontal/prefrontal systems involved in polysensory integration (potentially supporting improved awareness and higher-order cognition).^3^ The subparafascicular extension of the sweet spot may reflect an added benefit of modulating the dense projections of this nucleus to the brainstem, beyond the Pf’s striatal and cortical connections.^23^ Alternatively, this ventral emphasis may point to the importance of modulating structures below the thalamus, like the midbrain, a potentially effective target in its own right.^8, 17^

Our results differ from recent studies of thalamic DBS for patients with TBI, which targeted the ‘wing’ of the central lateral (CL) nucleus,^13, 76^ specifically the medial dorsal tegmental tract (DTTm). Like the CM-Pf, the CL is similarly thought to play a key role in arousal regulation via its striatal and frontal connections.^77^ Experimental studies in non-human primates show that CL stimulation can facilitate task performance^78^ and awaken animals from anesthesia.^79^ Following an earlier case study,^13^ a recent randomized trial in 6 patients with TBI—none with DoC—found improvement in executive function after CL/DTTm DBS.^76^

Direct comparisons of CL and CM-Pf stimulation are limited. One study found that stimulation of the CL/DTTm, but not the CM-Pf, improved behavioral performance in macaques, with the authors suggesting that CL-specific improvements may stem from its selective projections to striatal medium spiny neurons and broad effects on the frontal lobe,^78^ unlike the CM-Pf’s more variable striatal/cortical connections (as discussed above). However, another possibility is that therapeutic effects of these two targets converge not at the level of the CL and CM-Pf complex *per se*, but at the level of the CL and Pf specifically—perhaps, via a shared influence upon the dorsal striatum and associative and limbic regions of frontal and anterior cingulate cortex,^80, 81^ unlike the CM’s pattern of sensorimotor connections.^22, 23^

Stimulation was most beneficial when delivered to white matter pathways connecting the thalamus to the brainstem, hypothalamus, cerebellar dentate nucleus, and, to a lesser degree, the medial frontal cortex. This distribution which is thought to sustain resting wakeful states and connects to several cortical networks, particularly ones anchored in frontal and parietal cortex involved in awareness and cognition, including the default-mode, frontoparietal control, and dorsal attention networks.^82^ The dAAN is thought to dynamically interact with these cortical networks to provide a neuroanatomic basis linking the two foundational components of human consciousness: arousal and awareness.^40^

The pattern of optimal connectivity shows intriguing similarities to the neural circuitry underlying circadian regulation of arousal,^83^ which has been implicated in the pathophysiology and recovery of DoC.^84, 85^ Orexin-expressing neurons in the dorsomedial hypothalamus send dense projections to the locus coeruleus, the main site for norepinephrine synthesis, which has widespread excitatory effects.^83^ Activity in the locus coeruleus shows circadian variation, promoting arousal during wakefulness and being inhibited during sleep, and is under direct control by projections from the hypothalamus.^83^ Hypothalamic lesions disrupt this rhythm, causing somnolence, altered body temperature, and coma-like states.^46, 86^ Patients with DoC similarly show abnormal daily rhythms in EEG, temperature, and hormones.^84, 85^ The strength of circadian variation correlates with DoC severity, predicts recovery, and may even have therapeutic effects when exogenously entrained via, for instance, bright light stimulation.^84, 85^ These findings may relate to the integrity of neural circuits driving these circadian rhythms, consistent with our observation of enhanced connectivity from DBS sites to the hypothalamus and locus coeruleus in patients who improved.

Our findings may aid with understanding pathophysiology, predicting outcomes, and developing new treatments, both for DoC and other conditions. Brain areas showing stronger connectivity with effective DBS sites overlapped with the networks underlying acute-onset lesions causing arousal impairments^45, 46^ and epileptiform events associated with transient lapses of awareness.^48–50^ This suggests that effective stimulation targets for DoC might have similar benefits for a broader landscape of consciousness-impairing conditions. The findings may also aid with selecting cortical targets for non-invasive therapies like transcranial magnetic stimulation or transcranial direct current stimulation.^21^

The choice of stimulation paradigm is an important consideration when comparing our findings to prior work. We used a “medium” stimulation frequency of 20-40 Hz, motivated by positive results in earlier studies,^8, 18^ while others used higher frequencies of 100-185 Hz,^10, 13, 76^ likely having divergent effects.^87^ The optimal paradigm for DoC, and whether it differs by target, brain state, or species, is uncertain. Several authors^78, 79, 88, 89^ have hypothesized that stimulation efficacy may partially depend on the intent of DBS—e.g., awakening from anesthesia or sleep versus enhancing arousal in an already conscious subject—as well as correspondence between the stimulation frequency and resonant (intrinsic) frequencies of the intended brain state. For example, 150-225 Hz stimulation of the CL facilitated task performance in awake macaques,^89^ echoing the high frequency of excitatory input required to trigger dendritic electrogenesis in neocortical neurons.^89^ In contrast, another study found that CL stimulation at a lower (50 Hz), but not higher (200 Hz), frequency was effective at rousing macaques from propofol/isoflurane-induced anesthesia,^79^ perhaps reflecting entrainment of central thalamic neurons that fire at similar frequencies (20-40 Hz) during wakeful states.^90^ Conversely, low frequency (10 Hz) optogenetic stimulation of the central thalamus elicited spindle-like oscillations and behavioral arrest in rats,^88^ resembling patterns seen at the onset of sleep.^91^

Hence, a possible reason for the efficacy of 20-40 Hz DBS seen here could be the alignment with thalamic oscillations during wakefulness—akin to its rousing effects in anesthetized subjects.^79^ Consistent with this, Arnts et al. recently reported that 30-50 Hz DBS of the CM-Pf produced stronger treatment effects than 130 Hz stimulation in one patient with DoC^19^ and another with akinetic mutism.^92^ However, reports of contrary findings (e.g., restoration of consciousness during 130-180 Hz DBS in anesthetized macaques)^93, 94^ and potential interactions with other stimulation parameters (e.g., amplitude)^78, 89, 94^ highlight the need for further work to elucidate optimal paradigms and treatment mechanisms.

This study has limitations due to its clinical and retrospective nature. Patients had various DBS device models, with subtle differences in stimulation parameters, though not significantly associated with outcome. As in previous studies,^10, 11, 15, 18^, may have contributed to improvement in some patients; however, the lag time to DBS did not significantly differ between improved and non-improved groups, arguing against this as a major confound. Patients showed structural brain abnormalities, including cortical injuries and diffuse atrophy, presenting challenges for accurate image registration and raising questions about using normative data^37, 95^ to assess connectivity in patient brains.^35, 96^ We mitigated this by excluding patients with severe abnormalities preventing accurate template alignment and assessed the predictive utility of our findings using cross-validation techniques and examining relevance to patients with consciousness-impairing lesions^45, 46^ and seizures.^48, 49^ However, future replications will be important.

## Data Availability

Anonymized patient-level clinical information is available in Supplementary Material. The optimal stimulation site (sweet spot) and structural connectivity results will be available within Lead-DBS software upon publication (www.lead-dbs.org). Normative functional MRI and diffusion MRI data are publicly available:
https://datadryad.org/stash/dataset/doi:10.5061/dryad.nzs7h44q2
https://dataverse.harvard.edu/dataset.xhtml?persistentId=doi:10.7910/DVN/25833

## ACKNOWLEDGEMENTS

We thank Charles Jennings for his advice and helpful comments during revision of the manuscript.

## DATA AVAILABILITY

Anonymized patient-level clinical information is available in Supplementary Material. The optimal stimulation site (“sweet spot”) and structural connectivity results will be available within Lead-DBS software upon publication (www.lead-dbs.org). Normative functional MRI and diffusion MRI data are publicly available: https://datadryad.org/stash/dataset/doi:10.5061/dryad.nzs7h44q2 https://dataverse.harvard.edu/dataset.xhtml?persistentId=doi:10.7910/DVN/25833

## CODE AVAILABILITY

Code used to analyze the dataset is openly available within Lead-DBS software (https://github.com/leaddbs/leaddbs).

## METHODS

### Methods overview: a pragmatic analysis approach

This cohort is the largest sample of patients with DoC undergoing DBS reported to date, making it a valuable dataset to address key scientific and clinical questions. However, this patient population presents unique challenges, including abnormal neuroanatomy^1–3^ and low DBS response rates (∼30% in previous studies^9, 12, 18, 97^), making some analysis conventions unsuited to such a rare population. Given these challenges, we made pragmatic decisions in our analysis and reporting.

Accurate brain alignment to a common template is required for group-level analysis of optimal DBS locations and connectivity.^35^ We excluded patients with severe brain abnormalities causing poor template alignment, as determined by two neuroimaging experts blinded to clinical outcomes. However, these patients were retained for other analyses not requiring template alignment, including clinical variables associated with improvement. For clarity, **Supplementary Material** details the specific analyses each patient’s data contributed to, and the sample size for each analysis is noted throughout the results section.

Patients did not undergo advanced MRI connectivity sequences like functional or diffusion MRI. For analysis of optimal connectivity profiles, we used normative connectivity data acquired in healthy participants, an approach we have previously used to generate robust predictive models of DBS outcome in Alzheimer’s disease,^38^ epilepsy,^98^ Parkinson’s disease,^99^ and more.^35^

Finally, for hypothesis tests involving multiple comparisons, we report both uncorrected and false discovery-rate (FDR)-corrected *p*-values. This approach ensures transparency while balancing statistical rigor with the importance of preserving the exploratory insights afforded by this unique dataset.

### Study design and ethics

This was a retrospective analysis of a previous clinical study^9, 12^ of patients with DoC who underwent DBS at Dubrava University Hospital in Zagreb, Croatia. The University Hospital and the School of Medicine at the University of Zagreb, and informed consent was obtained from patients’ families or caregivers. Approval for retrospective data analysis performed in the current study was received from the Mass General Brigham institutional review board.

### Patients and DBS surgery

The analysis cohort included 40 patients. A description of study procedures and outcomes from 32 of these patients has been previously published.^9, 12^ Patients were selected based on neurophysiologic, clinical, and neuroimaging evaluations.^12^ Briefly, inclusion criteria included: (i) meeting clinical diagnostic criteria for UWS or MCS;^100, 101^ (ii) a minimum DoC diagnosis duration of 6 weeks; (iii) obtainable somatosensory evoked potentials (SSEP) via median nerve stimulation, with or without SSEPs from tibial nerve stimulation; (iv) periods of desynchronized scalp EEG activity observed during 12-24 hours of monitoring; (v) sufficient hemodynamic and respiratory stability to undergo study procedures; and (vi) absence of significant lesions (e.g., hemorrhages or infarctions) in the brainstem, diencephalon, or basal ganglia.^12^ The last criterion was based on the hypothesis that recovery potential depends on the integrity of subcortical nuclei and their dynamic interactions with cortical networks.^3^ For MRI examples of patients who met and did not meet this criterion, see figure 1 in Chudy et al.^12^.

The DBS procedure involved unilateral implantation of the CM-Pf. The rationale for unilateral, as opposed to bilateral, implantations was based on experience in prior studies, and to reduce surgical risk.^8, 18^ Most cases (37/40 patients) were implanted on the left (typically dominant) hemisphere, also motivated by experience in earlier studies^8, 18^ The remaining 3/40 were implanted on the right due to left-sided injuries or anatomical variations that made the right thalamus a more surgically feasible target. For group-level analyses, we flipped these right DBS leads to the left hemisphere by mirroring patients’ MRI/CT scans about the *x* axis as a first step, prior to further image processing steps.^35^

Device models varied and included Medtronic lead models 3387 or 3389, as well with outcomes (**Table 1**). Surgical target coordinates were defined on preoperative CT using the Schaltenbrand-Bailey^102^ atlas: 4.5 mm anterior to the posterior commissure, 1 mm inferior to the inter-commissural line, and 4 mm lateral to the third ventricular wall. The lateral coordinate was defined with respect to the third ventricular wall (rather than the inter-commissural line) to account for widened ventricles due to brain atrophy typically seen in this patient group.^9, 103^

Three days post-surgery, DBS devices were programmed to deliver a stimulation paradigm optimized per patient to elicit the strongest arousal reaction, as previously described.^9, 12^ Briefly, this involved testing each electrode contact using a stimulation frequency of 20-40 Hz, pulse width of 120-330 μs, and amplitude of 2-4.5 V or 2.5-5.5 mA. An arousal reaction was defined by eye opening (if the patient’s eyes were closed) with mydriasis and change in facial expression, with or without head turning and elevation of blood pressure and heart rate.^9, 12^ Stimulation parameters for each patient are reported in **Supplementary Material.** Stimulation was administered for 30 minutes every 2 hours during the day and ceased at night with the aim of promoting circadian (sleep-wake) cycles.^8^

### Clinical outcomes

Outcomes were tracked using the Coma Recovery Scale-Revised (CRS-R).^26^ Total scores range from 0-23, with higher scores indicating a higher level of consciousness across auditory, visual, motor, oromotor, communication, and arousal subscales.^26^ Supplementary clinical measures included the Disability Rating Scale (DRS) and Coma/Near-Coma (C/NC) scale.^27^ Patients were classified as having unresponsive wakefulness syndrome (UWS), a minimally conscious state (MCS), or full consciousness.^1^ We defined “improved” patients as those who transitioned from UWS to MCS or conscious, or from MCS to conscious, within 12 months post-DBS, while “non-improved” patients were those who did not change states, as in our previous work.^9, 12^ Dichotomization of the cohort into improved and non-improved groups was intended to enhance our sensitivity to factors driving clinically significant improvements in consciousness.^104^ However, we performed secondary analyses using each patient’s change in CRS-R scores to test consistency across a different definition of improvement.

### DBS electrode localization and stimulation-induced electric fields

DBS electrodes were reconstructed using Lead-DBS software (**Fig. 1**).^35^ Given the study’s retrospective nature, the types of pre- and post-operative imaging data varied (MRI, CT, or both). Electrode localizations were therefore optimized per patient, based on available data. When a T1 MRI scan was available (28/40 patients), it was used as the reference image for non-linear spatial warping to template space; in other cases, we used CT. MRI scans were acquired on a 1.5 T Siemens Avanto or Aera scanner using a volumetric T1-weighted magnetization-prepared rapid acquisition gradient echo (MPRAGE) sequence with a voxel resolution ≤1mm^3^. CT scans were acquired using a Siemens scanner with slice thickness ≤0.5mm.

We first linearly co-registered the post- to the pre-operative image using Advanced Normalization Tools software,^105^ then calculated nonlinear spatial warps to the Montreal Neurological Institute (MNI) 152 ICBM 2009b nonlinear asymmetric brain template. To accommodate the heterogenous imaging modalities (MRI or CT) available for these nonlinear warps, we used a recently developed deep learning-based tool, EasyReg,^106^ which can perform robust, modality-agnostic registrations,^107^ unlike classical techniques that rely upon optimization of similarity metrics between images.^106^As shown in Billot et al.,^33^ this strategy can cope with CT scans, despite their low soft-tissue contrast. This is because synthetic images with low contrast-to-noise ratio are regularly seen during model training.^106^ To further optimize the performance of EasyReg for CT, we followed the approach of Billot et al.,^33^ and stretched the histogram of CT values in the soft-tissue interval (0<HU<80) using the piece-wise linear “tone-mapping” function implemented in Lead-DBS software.^35^

Accuracy of image registrations was reviewed by two authors (AELW and AH), blinded to clinical outcomes. Twelve patients were excluded from further analysis involving DBS localizations, primarily due to severe brain atrophy and/or grossly enlarged ventricles resulting in poor template alignment. However, these patients were retained for other analyses not requiring alignment to MNI space. Details of the specific patients included in each analysis are provided in **Supplementary Material**.

DBS electrodes were localized using the PaCER^108^ or TRAC/CORE^35^ algorithm in Lead-DBS.^35^ We then calculated electric fields (E-fields) for each patient’s stimulation settings using the finite element method (FEM) implemented in FieldTrip/SimBio.^36^ E-fields represent the first derivative of the estimated voltage distribution applied to voxels in space; the field’s magnitude is strongest near active electrode contacts and diminishes rapidly with distance.^38^

### Analysis of optimal brain tissue integrity

We assessed whether whole-brain and subcortical tissue volumes differed between improved (*n=*8) and non-improved (*n=*18) groups. Each patient’s T1- weighted MRI scan was segmented into whole-brain volumes of gray matter, white matter, and cerebrospinal fluid using SynthSeg software.^33^ Additionally, we segmented regional subcortical volumes and carefully inspected the results for accuracy. To adjust for inter-patient variability in brain size, each segmented volume was normalized by the total intracranial volume. We additionally normalized by tissue volumes from age-matched samples of T1-weighted MRI scans from the Nathan Kline Institute-Rockland Sample (NKI-RS).^34^ This longitudinal, community-ascertained neuroimaging study includes >1,500 individuals aged 6-85 years. For each DoC patient, we selected a subset of T1- weighted MRI scans from NKI-RS control participants whose ages matched the patient’s age within a ±2-year range. On average, we found 92 matching control participants per patient (range=46-126). The NKI-RS scans were processed using SynthSeg,^33^ normalized by total intracranial volume and used to convert each patient’s whole-brain and subcortical volumes into *z*-scores.

### Analysis of optimal stimulation sites

To identify optimal stimulation sites, we compared E-fields between improved (*n=*10) and non-improved (*n=*18) groups.^35^ As in prior studies,^38, 39^ we focused on assumed estimate of the voltage required to activate axons.^38, 109^ We initially conducted a series of voxel-wise *t-*tests, resulting in a map of *t-*scores (*t-*map) where positive values indicate higher E-field magnitudes in the improved relative to non-improved patients. To identify a candidate site of optimal stimulation, the *t*- map was thresholded at *p*<0.05 (uncorrected). We then subjected this candidate site to *k-*fold cross-validation with *k=*10, where *k* was the number of groups into which the dataset was randomly split.^35, 38^ A *k* of 10 was used to align with previous similar DBS studies.^38, 99^ We iteratively used *k-*1 folds for training and the remaining fold for testing. In each iteration, the *t*-map was recalculated, leaving out the E-fields of patients in the test fold. The clinical outcomes for the left-out patients were then estimated by calculating the peak value of a voxel-wise multiplication of their E-field distributions with the derived *t-*map. These estimates were then compared between the improved and non-improved patients using a two-sample *t*-test. The intuitive interpretation of this analysis is that positive values in the *t*-map represent better stimulation locations. By testing whether E-fields from left-out patients more strongly overlapped with the positive sites in the *t-*map, we evaluated the robustness and potential predictive utility of our findings.^35, 38^

### Analysis of optimal structural connectivity

To identify white matter tracts associated with improvement, we utilized the fiber filtering approach in Lead-DBS software with normative structural connectome (**Fig. 4**)^35, 38^. Given the potential importance of small and intricate connections within and around the thalamus, and to the brainstem, we used a state-of-the-art, ultra-high resolution (760 µm) diffusion-weighted MRI dataset acquired across 18 hours,^37^ as detailed in our recent work.^38, 39^ Like the analysis of optimal stimulation sites, which analyzed stimulations across voxels, here we examined stimulations across *streamlines* of the normative connectome in the same, mass-univariate fashion. For each streamline and E-field pair, we recorded the peak magnitude that the streamline traversed. Then, we performed the same *t*-tests on the E-field magnitudes between improved and non-improved groups, yielding a *t*-value for each streamline, with positive *t*-scores indicating exposure to E-fields that were higher in the improved group. To identify a candidate network of optimal structural robustness of this network using the same *k-*fold cross-validation (*k=*10) procedure. Specifically, we iteratively assigned *t-*scores to streamlines, each time leaving out E-fields of patients in the test fold. We then computed the peak overlap between the left-out E-fields and the t-weighted streamlines for each patient, comparing the results between groups using a two-sample t-test.^35, 38^ To define subcortical nuclei traversed by the observed fiber tracts, we compared results to atlases of the hypothalamus^41^ (https://zenodo.org/records/3942115), brainstem (https://www.nitrc.org/projects/brainstemnavig), ascending arousal network^40^ (https://doi.org/10.5061/dryad.zw3r228d2), and cerebellum^42^ (https://www.diedrichsenlab.org/imaging/propatlas.htm).

### Analysis of optimal functional connectivity

We investigated blood-oxygen-level-dependent (BOLD) connectivity of DBS sites using a normative, sex-balanced sample of resting-state fMRI scans acquired in 1,000 healthy adults (500 males, 500 females) from the Brain Genomics Superstruct Project.^43, 44^ The fMRI data and pre-processing pipeline are publicly available.^95^ For each patient’s E-field location, we calculated the mean BOLD time-course (in the normative scans) using a weighted average across all voxels with E-field magnitudes >200 V/m, then measured connectivity with every brain voxel using Fisher’s *r-*to-*z* transformed Pearson correlations. Connectivity strengths were then compared between improved and non-improved groups using voxel-wise *t-*tests in permutation analysis of linear models (PALM) software.^110^ The results of this analysis was a spatially continuous (i.e., unthresholded) map of brain areas showing greater functional coupling with DBS sites linked to improvement (positive *t-*scores) or non-improvement (negative *t-*scores).

### Comparison with consciousness-impairing lesion network

We hypothesized that the brain network underlying DBS improvement in DoC may overlap with the pathological circuits underlying consciousness-impairing brain lesions. In other words, we reasoned that the network where stimulation improves consciousness may reflect the network where lesions disrupt it. To test this, we current study—who had acute-onset lesions in the thalamus or brainstem due to stroke or head trauma. The patients were obtained from two sources: one study of patients with lesion-induced coma,^45^ and another of patients with variable outcomes ranging from coma to no impairment (i.e., awake).^46^ In the latter study, patients were ordinally ranked using scores from 1-6,^46^ based on clinical definitions of Plum and Posner,^111^ with lower scores indicating more severe impairment (coma=1; stupor=2; obtunded=3; somnolent=4; lethargic=5; awake=6). We combined the two datasets by assigning all coma patients a score of 1 (in both studies) while retaining the original rankings from the second study for patients with outcomes less severe than coma (i.e., scores from 2-6). Using the same normative resting-state fMRI data^43, 44, 95^ and processes described earlier, we used binary lesion masks as seeds and calculated functional connectivity with all brain voxels to create a lesion connectivity map for each patient. We then calculated a similarity score between each patient’s lesion connectivity map and our DBS improvement network (**Fig. 5A**) using spatial (Pearson) correlations. Finally, we tested whether higher similarity to our DBS improvement network was associated with worse outcomes (i.e., lower arousal scores) using a rank-based, non-parametric Spearman correlation (**Fig. 5B**).

Since the latter study^46^ included both patients with coma and others with more variable levels of impairment, we conducted an additional analysis focusing solely on patients with lesion-induced coma (*n=*14) and those who were awake (*n=*15), comparing the groups using a two-sample *t-*test.

### Comparison with consciousness-impairing seizure network

In a final analysis, we explored whether effective DBS sites for DoC modulate the same network that is disrupted by absence seizures, which are brief lapses of awareness marked by generalized spike-wave discharges (GSW) on scalp EEG. We used findings from a previous study of 15 patients with absence epilepsy who underwent up to 60 mins of EEG-fMRI.^48–50^ GSW timings were manually marked on the EEG and used as regressors in a whole-brain fMRI analysis to identify discharge-related BOLD signal changes. Event-related independent component canonical hemodynamic response function (HRF), which is often seen with epileptiform events.^113, 114^ The eICA was performed on temporally concatenated fMRI data from all patients, covering a 32-second window before and after GSW onset.^48, 112^ Thirteen brain components significantly associated with GSW were identified (*F-*test; *p*<0.05, Bonferroni-corrected), each represented by a spatial map (*z-*scores) and a BOLD time-course. Positive *z-*scores indicated regions with increased BOLD signal (activation) and negative *z-*scores indicated decreased signal (deactivation). We averaged all *z-*score maps together to create one map representing overall patterns of activation/deactivation (**Fig. 5C**). Finally, both this map and our DBS improvement network were warped to FreeSurfer’s *fsaverage5* template.^115^ Spatial similarity was then measured using a Spearman correlation, with significance assessed via spin-permutation testing (10,000 spins).^51^

### Statistical analyses

Analyses were performed using MATLAB version R2023b and RStudio version 2022.07.01. For comparison of clinical variables and MRI tissue volumes between improved and non-improved groups, we used non-parametric, permutation-based two-sample *t-*tests (10,000 permutations) for continuous variables and Fisher’s exact tests for categorical variables. Statistical procedures for the remaining analyses are described in the methods. Significance was defined using an alpha of 0.05 (two-tailed). For hypothesis tests involving multiple comparisons, we report both uncorrected and Benjamini-Hochberg^116^ FDR-corrected *p-*values.

