## Supplementary Material for "A human brain network linked to restoration of consciousness after deep brain stimulation"

**Joint senior authors*

**Correspondence**

Aaron E.L. Warren

Department of Neurosurgery, Mass General Brigham, Harvard Medical School

Hale Building for Transformative Medicine, 60 Fenwood Road, Boston, MA, USA

**Contents**

**S1:** Anonymized patient-level data

**S2:** Secondary analyses

**S3:** Supplementary references

**S1: Anonymized patient-level data**

Detailed patient-level information, including DBS settings and coordinates of stimulation locations in MNI 152 ICBM 2009b nonlinear asymmetric template space, is available in Excel spreadsheet format on the open science framework (OSF):

*Link provided upon publication.*

**S2: Secondary analyses**

We repeated our analyses across three secondary analysis designs: (i) using a finer outcome measure based on the change in total CRS-R scores (instead of a binary improved/non-improved classification), (ii) excluding three patients who died within 12 months of DBS, and (iii) excluding three patients with right-sided DBS implants.

For associations between CRS-R increases and clinical and demographic variables and MRI brain tissue volumes, we fit linear models using the *lm* function in RStudio version 2022.07.01. For the optimal stimulation site and structural connectivity analyses, E-field magnitudes were correlated against CRS-R score increases using Spearman correlations.[1] All other procedures the same as those reported for the primary analyses in the main text.

S2 Tables 1-3 report associations between outcomes and clinical and demographic variables. S2 Figures 1-3 report associations with MRI tissue volumes. S2 Table 1 reports optimal stimulation site results across primary and secondary analyses. S2 Figure 4 reports optimal structural connectivity results across primary and secondary analyses.

| **Demographic and clinical variables** | **Association with change in CRS-R scores after DBS** ***(F-stat. [df], uncorrected p-value [FDR-corrected], R-squared, β)*** |
|---|---|
| Age at injury (years) | F(1,38)=8.38, p=0.006 (0.04), R-squared=0.18, β=-0.13 |
| Sex (male or female) | F(1,38)=0.32, p=0.58 (0.68), R-squared=0.008 |
| Time from injury to DBS (months) | F(1,38)=1.61, p=0.21 (0.56), R-squared=0.04 |
| C/NC score before DBS | F(1,38)=9.72, p=0.003 (0.04), R-squared=0.20, β=-2.3 |
| DRS score before DBS | F(1,38)=6.18, p=0.02 (0.09), R-squared=0.14, β=-2.5 |
| CRS-R score before DBS | F(1,38)=0.89, p=0.35 (0.67), R-squared=0.02 |
| DoC state prior to DBS (UWS or MCS) | F(1,38)=1.54, p=0.22 (0.56), R-squared=0.04 |
| Implant side (left or right) | F(1,38)=0.80, p=0.38 (0.67), R-squared=0.02 |
| Cause of injury (CA or TBI) | F(1,38)=0.0007, p=0.98 (0.98), R-squared=1.881e-05 |
| DBS lead model (M3387, M3389, or BSCI) | F(2,37)=0.24, p=0.79 (0.85), R-squared=0.01 |
| Stimulation amplitude (V)† | F(1,24)=0.35, p=0.56 (0.68), R-squared=0.01 |
| Stimulation amplitude (mA)† | F(1,12)=0.37, p=0.55 (0.68), R-squared=0.03 |
| Stimulation frequency (Hz) | F(1,38)=0.45, p=0.51 (0.68), R-squared=0.01 |
| Stimulation pulse width (μs) | F(1,38)=1.44, p=0.24 (0.56), R-squared=0.04 |

**S2 Table 1: Associations between increases in CRS-R scores and demographic and clinical variables (*n*=40).** Associations were performed by fitting linear models with each patient's increase in CRS-R score as the dependent variable. Beta coefficients (*β*) are provided for significant associations (*p*<0.05, uncorrected), indicating the direction of the association (positive or negative).

*<u>Abbreviations</u>:* BSCI, Boston Scientific Vercise lead model; CA, Cardiac arrest; CNC, Coma/Near-Coma; CRS-R, Coma Recovery Scale-Revised; DRS, Disability Rating Scale; M3387, Medtronic lead model 3387; M3389, Medtronic lead model 3389; MCS, Minimally conscious state; OR, Odds Ratio; TBI, Traumatic brain injury; UWS, Unresponsive wakefulness syndrome.

†*Note: associations with stimulation amplitude were performed separately in patient sub-groups for whom amplitude was recorded as voltage (V; n=26) or milliamps (mA; n=14).*

| | DoC improved (*n*=10), median (IQR) or proportions | DoC non-improved (*n*=27), median (IQR) or proportions | *p*-value uncorrected (FDR-corrected), effect size (95% CI) |
|---|---|---|---|
| Age at injury (years) | 19 (16-33) | 39 (24-53) | *p*=0.05 (0.23) Hedge's *g*=-0.7 (-1.62, 0) |
| Sex (Male:Female) | 7:3 | 21:6 | *p*=0.7 (0.92), OR=0.67 (0.1, 5.28) |
| Time from injury to DBS (months) | 8 (3-12) | 6 (3-15) | *p*=0.69 (0.92), Hedge's *g*=-0.19 (-0.53, 0.61) |
| C/NC score before DBS | 2 (1-2) | 3 (2-3) | *p*=0.006 (0.07), Hedge's *g*=-1.12 (-2.06, -0.46) |
| DRS score before DBS | 2.3 (1.8-2.6) | 3.2 (2.6-3.4) | *p*=0.01 (0.07), Hedge's *g*=-1.0 (-2.23, -0.35) |
| CRS-R score before DBS | 7 (6-8) | 4 (4-6) | *p*=0.11 (0.39), Hedge's *g*=0.59 (-0.02, 2.25) |
| DoC state prior to DBS (UWS:MCS) | 7:3 | 24:3 | *p*=0.31 (0.72), OR=0.3 (0.03, 2.78) |
| Implant side (Left:Right) | 9:1 | 25:2 | *p*=1.0 (1.0), OR=0.73 (0.03, 47.14) |
| Cause of injury (CA:TBI) | 7:3 | 18:9 | *p*=1.0 (1.0), OR=1.16 (0.2, 8.63) |
| DBS lead model (M3387:M3389:BSCI) | 4:2:4 | 9:10:8 | *p*=0.72 (0.92) |
| Stimulation amplitude (V)$^{\dagger}$ | 3.25 (3-3.5) | 3 (2.3-3.5) | *p*=0.47 (0.84), Hedge's *g*=0.32 (-0.22, 0.91) |
| Stimulation amplitude (mA)$^{\dagger}$ | 4.25 (4-5) | 4.5 (4.25-5) | *p*=1.0 (1.0), Hedge's *g*=-0.11 (-2.04, 1.25) |
| Stimulation frequency (Hz) | 35 (25-40) | 30 (25-30) | *p*=0.26 (0.72), Hedge's *g*=0.44 (-0.39, 1.45) |
| Stimulation pulse width (µs) | 210 (210-210) | 210 (180-210) | *p*=0.48 (0.84), Hedge's *g*=0.28 (-0.15, 0.78) |

**S2 Table 2: Comparison of demographic and clinical variables between patients who improved (*n*=10) versus patients who did not improve (*n*=27) with DBS, excluding those who died <12 months after DBS.** $^{\dagger}$*Note: comparisons of stimulation amplitude were performed separately in patient sub-groups for whom amplitude was recorded as voltage (V; n=6 improved versus n=19 non-improved) or milliamps (mA; n=4 improved versus n=8 non-improved).*

| | DoC improved (*n*=10), median (IQR) or proportions | DoC non-improved (*n*=27), median (IQR) or proportions | *p*-value uncorrected (FDR-corrected), effect size (95% CI) |
|---|---|---|---|
| Age at injury (years) | 21 (16-33) | 39 (25-55) | *p*=0.046 (0.20), Hedge's *g*=-0.75 (-1.67, -0.03) |
| Sex (Male:Female) | 8:2 | 20:7 | *p*=1.0 (1.0) OR=1.4 (0.20, 16.51) |
| Time from injury to DBS (months) | 5 (3-11) | 6 (4-15) | *p*=0.64 (0.81) Hedge's *g*=-0.21 (-0.57, 0.54) |
| C/NC score before DBS | 1.5 (1-2) | 3 (2-3) | *p*=0.006 (0.046) Hedge's *g*=-1.16 (-2.13, -0.50) |
| DRS score before DBS | 2.1 (1.8-2.6) | 3.2 (2.6-3.4) | *p*=0.007 (0.046), Hedge's *g*=-1.06 (-2.29, -0.41) |
| CRS-R score before DBS | 7.5 (7-9) | 4 (4-6) | *p*=0.08 (0.26) Hedge's *g*=0.66 (0.03, 2.29) |
| DoC state prior to DBS (UWS:MCS) | 7:3 | 24:3 | *p*=0.31 (0.67) OR=0.3 (0.03, 2.78) |
| Cause of injury (CA:TBI) | 8:2 | 19:8 | *p*=0.69 (0.81) OR=1.66 (0.24, 19.48) |
| DBS lead model (M3387:M3389:BSCI) | 4:1:5 | 9:9:9 | *p*=0.42 (0.68) |
| Stimulation amplitude (V)† | 3.2 (3-3.4) | 3 (2.3-3.5) | *p*=0.65 (0.81) Hedge's *g*=0.23 (-0.31, 0.87) |
| Stimulation amplitude (mA)† | 4.5 (4-5) | 4.5 (4-5) | *p*=0.75 (0.81) Hedge's *g*=0.21 (-1.24, 1.06) |
| Stimulation frequency (Hz) | 35 (25-40) | 30 (25-30) | *p*=0.25 (0.65) Hedge's *g*=0.44 (-0.42, 1.44) |
| Stimulation pulse width (μs) | 210 (210-210) | 210 (180-210) | *p*=0.37 (0.68) Hedge's *g*=0.35 (-0.07, 0.84) |

**S2 Table 3: Comparison of demographic and clinical variables between patients who improved (*n*=10) versus patients who did not improve (*n*=27) with DBS, excluding those who underwent right-sided DBS implantations.** †*Note: comparisons of stimulation amplitude were performed separately in patient sub-groups for whom amplitude was recorded as voltage (V; n=5 improved versus n=18 non-improved) or milliamps (mA; n=5 improved versus n=9 non-improved).*

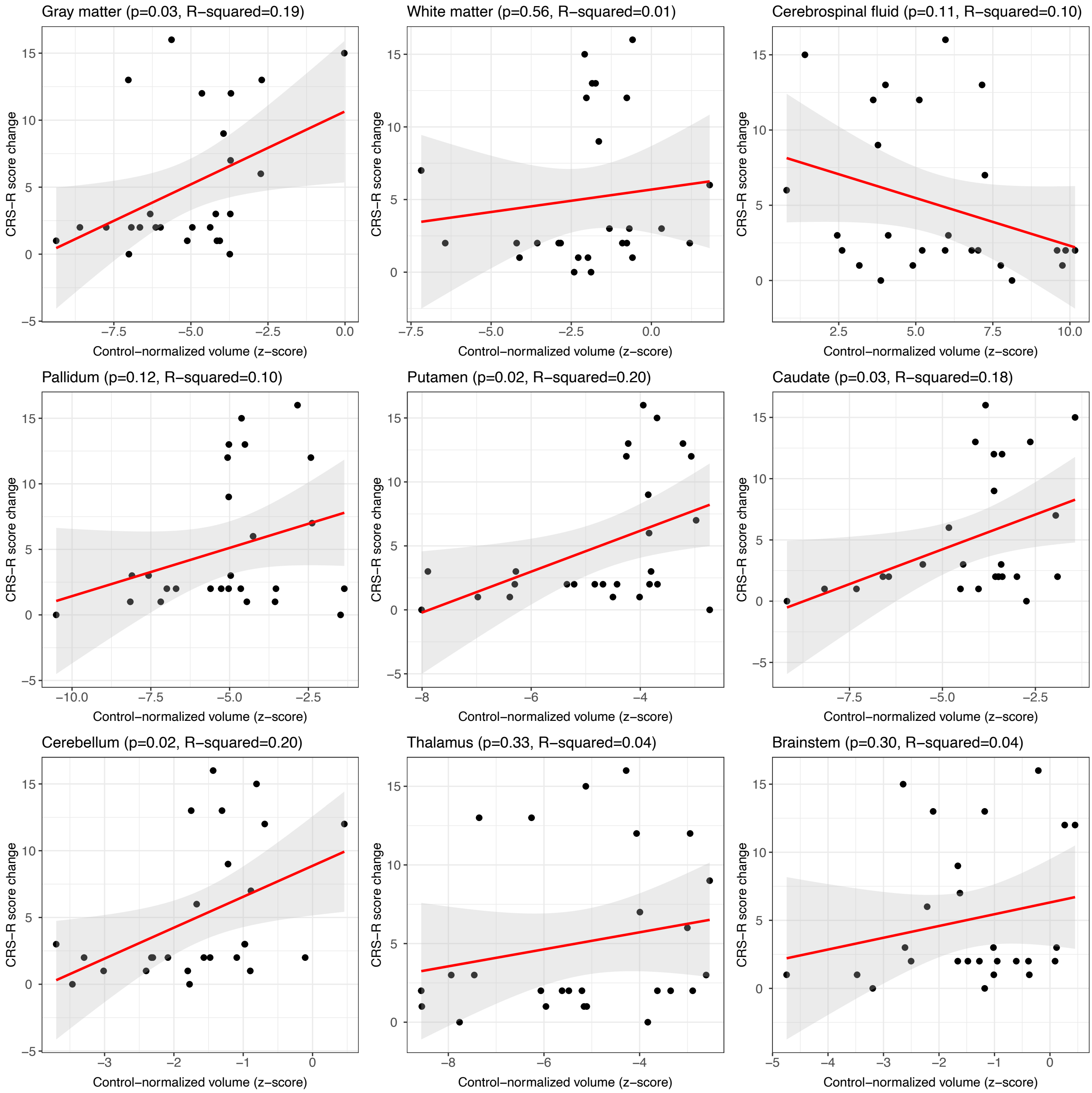


**S2 Figure 1: Associations between MRI tissue volumes and increases in CRS-R scores (*n*=26)**. Associations were performed by fitting linear models, with each patient's increase in CRS-R score as the dependent variable (*y* axis) and MRI tissue volume as the independent variable (*x* axis). Each patient's MRI tissue volumes were normalized by their total intracranial volume and then by the average value in age-matched controls from the Nathan Kline Institute-Rockland Sample (NKI-RS).[2] Volumes are expressed as z-scores measuring the distance, in units of SD, away from the control mean. * = $p<0.05$ (uncorrected).

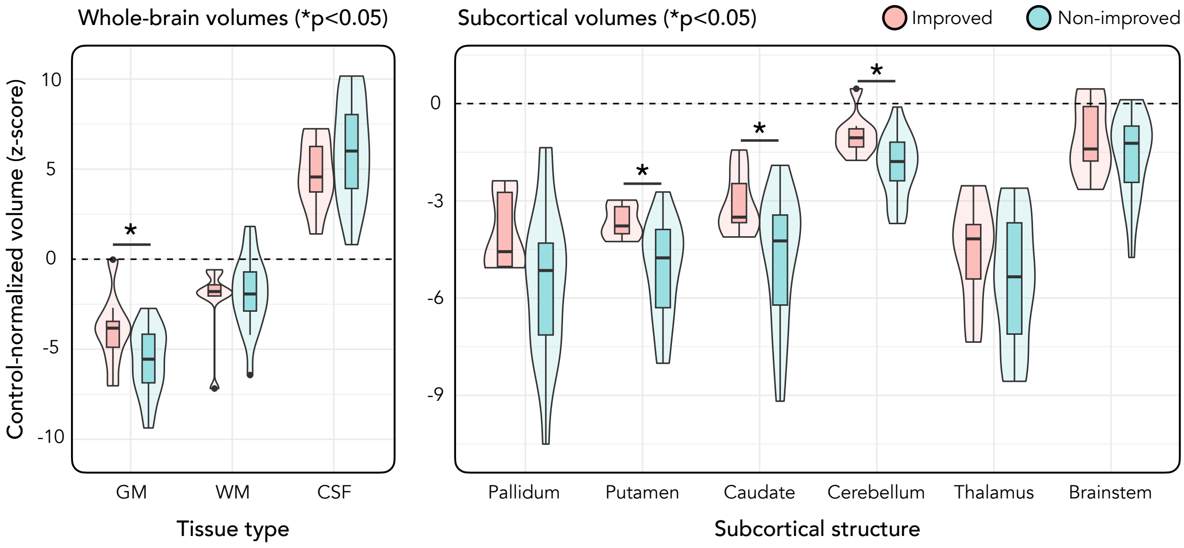


**S2 Figure 2: Comparison of MRI tissue volumes between improved (*n*=8) and non-improved (*n*=16) groups, excluding patients who died <12 months after DBS implantation**. Violin plots showing whole-brain volumes of gray matter (GM), white matter (WM), and cerebrospinal fluid (CSF) in the improved (pink) and non-improved (blue) groups. The plot on the right shows comparisons of subcortical gray matter volumes. The dashed horizonal line on each plot indicates the average value in age-matched controls from the Nathan Kline Institute-Rockland Sample (NKI-RS).[2] Volumes from patients are expressed as z-scores measuring the distance, in units of SD, away from the control mean. * = $p<0.05$ (uncorrected).

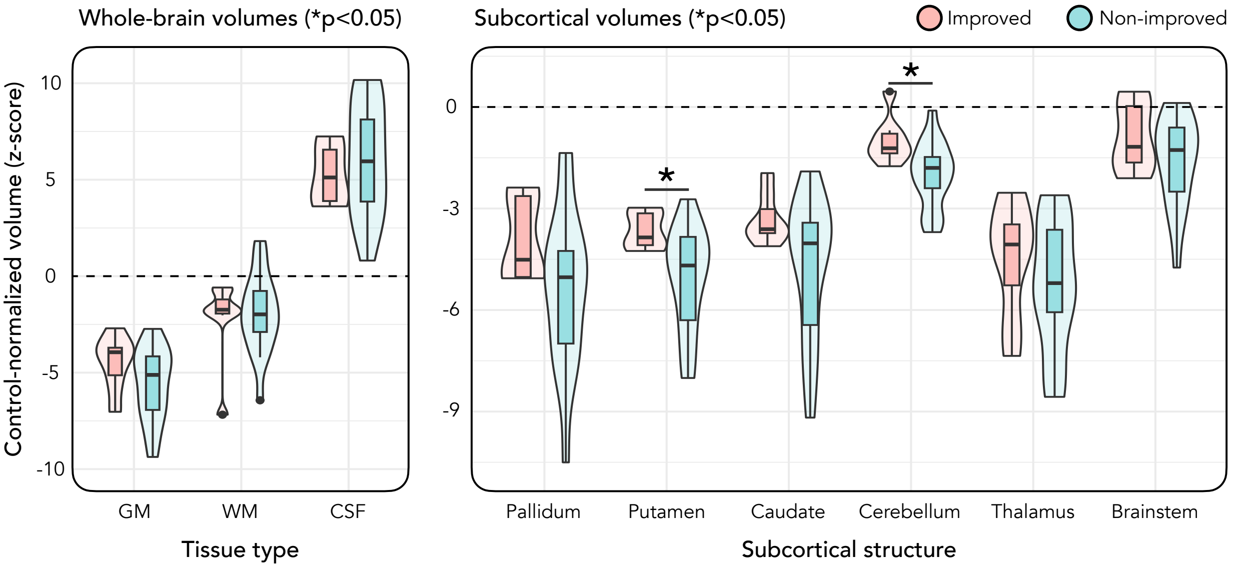


**S2 Figure 3: Comparison of MRI tissue volumes between improved (*n*=7) and non-improved (*n*=17) groups, excluding patients who underwent right-sided DBS implantations.** Violin plots showing whole-brain volumes of gray matter (GM), white matter (WM), and cerebrospinal fluid (CSF) in the improved (pink) and non-improved (blue) groups. The plot on the right shows comparisons of subcortical gray matter volumes. The dashed horizonal line on each plot indicates the average value in age-matched controls from the Nathan Kline Institute-Rockland Sample (NKI-RS).[2] Volumes from patients are expressed as z-scores measuring the distance, in units of SD, away from the control mean. * = $p<0.05$ (uncorrected).

| | **Coordinates (mm) for optimal stimulation site (*x, y, z;* MNI 152 2009b)** | **Euclidean distance (mm) from optimal stimulation site in primary analysis** |
|---|---|---|
| **Primary analysis** (*n*=10 improved vs *n*=18 non-improved) | -6.91, -20.11, -3.08 | 0 |
| **Secondary analysis 1: CRS-R scores** (*n*=28) | -7.1, -20.67, -3.44 | 0.69 |
| **Secondary analysis: excluding patients who died** (*n*=9 improved vs *n*=17 non-improved) | -6.99, -20.2, -3.42 | 0.36 |
| **Secondary analysis: excluding patients with right-sided DBS** (n=9 improved vs *n*=16 non-improved) | -6.71, -20, -3.08 | 0.24 |

**S2 Table 4: Coordinates of optimal stimulation sites for different analysis designs.** Analysis of optimal stimulation sites was repeated across four designs: the primary analysis reported in the main text, and three secondary analyses. In each analysis, the optimal stimulation site was defined as the center-of-gravity of the largest voxel cluster at a threshold of $p<0.05$ (uncorrected). Coordinates are provided as *x, y,* and *z* values (mm) in MNI 152 ICBM 2009b nonlinear asymmetric template space. For each site, the Euclidean distance (mm) away from the optimal stimulation site reported in the main text is also reported.

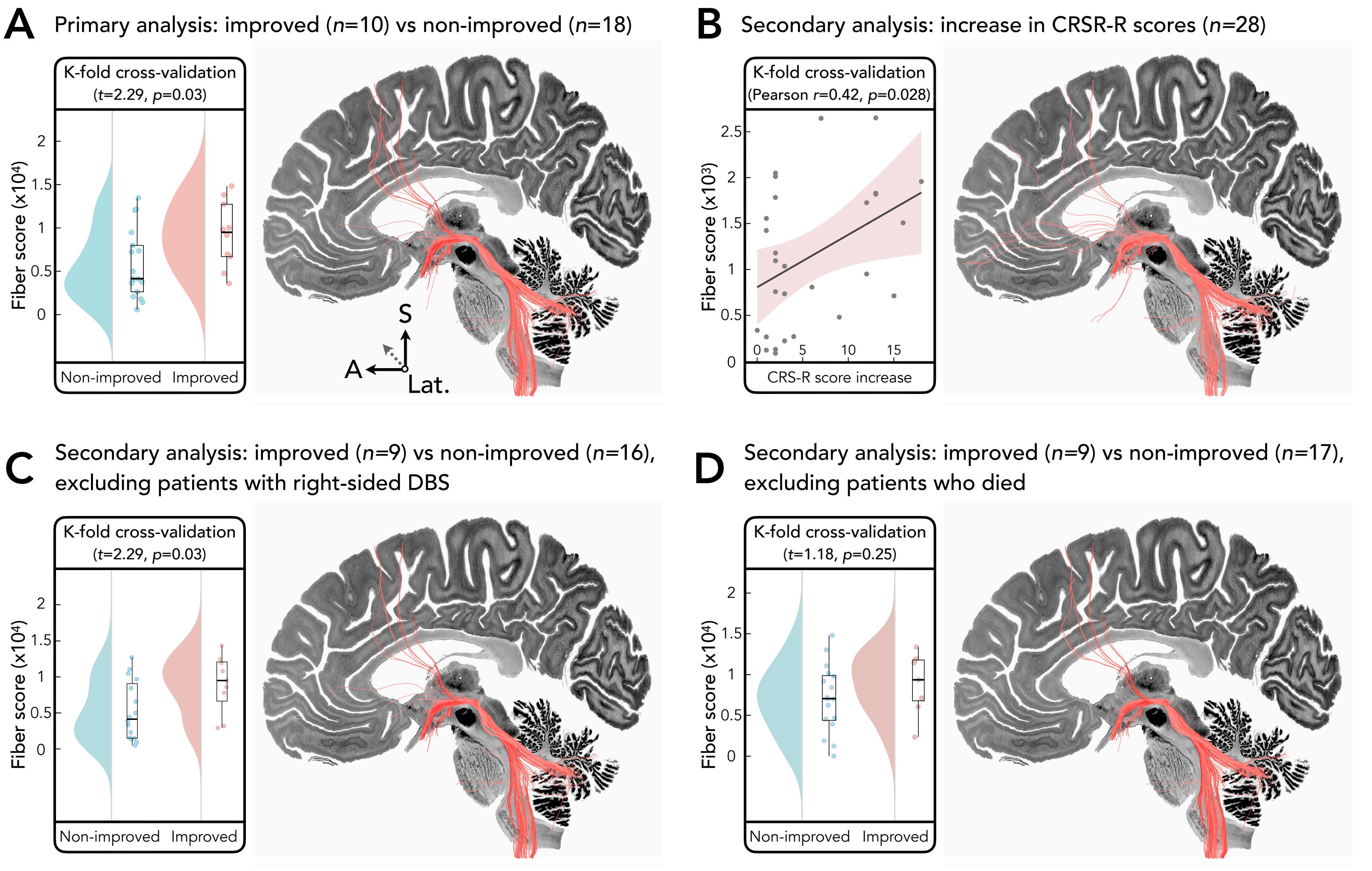


**S2 Figure 4: Optimal structural connectivity for different analysis designs.** Analysis of optimal structural connectivity was repeated across four designs: the primary analysis reported in the main text (A), and three secondary analyses (B-D). In each analysis, white matter fiber tracts associated with improvement are shown as pink-red lines ($p<0.05$, uncorrected). Box plots (A, C, D) or scatter plot (B) show the results of K-fold ($k=10$) cross-validation. Results are displayed upon the BigBrain histological atlas[3] registered to MNI space.[4]

*Abbreviations:* A, Anterior; CRS-R, Coma Recovery Scaled—Revised; Lat., Lateral; S, Superior.
